# Life beyond childhood: insight into the lived experience of 91 adults with KBG syndrome through an online patient/caregiver reported co-produced questionnaire

**DOI:** 10.1101/2024.11.19.24317531

**Authors:** KJ Low, M Walker, G Treneman-Evans, NC Bramswig, MK Herlin, G Lesca, E Scarano, CW Ockeloen, A Bayat

**Affiliations:** Centre for Academic Child Health, Bristol Medical School, University of Bristol, Bristol UK; Clinical Genetics, University Hospitals Bristol and Weston NHS trust, Bristol, UK; Avon and Wiltshire Mental Health Partnership NHS Trust, Adult ADHD service, Petherton Resource Centre, Bristol, UK; Centre of Medical Genetics, Department of Medical Genetics, University and University Hospital Münster, Münster, Germany; Department of Clinical Genetics, Aarhus University Hospital, Aarhus, Denmark; Department of Medical Genetics, Lyon University Hospitals, Claude Bernard Lyon 1 University, Lyon, France; Pediatric Unit, IRCCS Azienda Ospedaliero-Universiitaria di Bologna, Bologna, Italy; Department of Human Genetics, Radboud University Medical Center, Nijmegen, the Netherlands; Department of Pediatrics, Danish Epilepsy Center, Dianalund, Denmark; Department of Regional Health Research, University of Southern Denmark, Odense, Denmark

**Keywords:** ANKRD11, Adult, KBGS, Neurodevelopment

## Abstract

**Purpose:** KBG syndrome (KBGS) is a rare neurodevelopmental syndrome. We aimed to study the impact of KBGS in adulthood as reported by individuals with KBGS and their families/caregivers, thereby exploring aspects of everyday life under-reported by healthcare professionals.

**Methods:** We co-produced an online questionnaire for adults with KBGS and their families/caregivers. Participants were recruited via the KBG foundation, an American-based charity supporting individuals with KBGS and their families worldwide, and other international collaborators.

**Results:** There were 91 responses for analysis, across the age range of 16-86 years. Respondents described a range of living arrangements, education, employment, leisure activities and relationships. A higher proportion of 45-54year olds had achieved independent living skills such as driving and grocery shopping compared to the younger age groups. None of the participants who were experiencing seizures lived independently. We described high rates of psychiatric comorbidities, behavioural difficulties, sleep problems, seizures, visual and hearing problems, dental and skeletal issues, and a higher than expected cardiovascular and gastrointestinal burden of disease.

**Conclusion:** This study provides new insights into the everyday life of adults with KBGS, along with high rates of comorbidities which continue to impact on quality of life into adulthood, with implications for medical care.

## 1. Introduction

KBG syndrome (KBGS) (OMIM#148050), is a rare neurodevelopmental syndrome, caused by heterozygous pathogenic *ANKRD11* variants^1^. It is characterised by distinctive craniofacial features, macrodontia, short stature, skeletal abnormalities, intellectual and developmental disability, behavioural and psychiatric comorbidities and sometimes associated with seizures^1^. First described in 1975 and named after the surnames of the first families reported with the syndrome^2^, it is now known to affect more than 500 families’ worldwide (based on estimates from patient groups and published cohorts).

With the increasing availability of new genomic techniques and reducing test costs, more individuals are being diagnosed with KBGS. International paediatric cohorts have been described in the literature leading to a better understanding of the phenotypic spectrum in childhood^3^. However, when a child is diagnosed with KBGS, their parents are often concerned about long-term implications for their child, which also requires good knowledge of the adult phenotypic spectrum. Adults with syndromic neurodevelopmental disorders can experience fragmented care^4^, and there is a need for more data on the natural history of KBGS into adulthood in order to improve care planning and inform future guidelines.

We recently reported a study of 36 adults with KBGS based on clinician reported data^5^. The study presented here is the first to examine the long-term outcomes of KBGS into adulthood as reported by individuals with KBGS and/or their family members; with a particular focus on the impact of the disorder on daily life, employment, leisure and relationships. We co-produced a survey designed to enable individuals with KBGS, their families and physicians, to have a clearer picture of what they can expect for the future.

## 2. Methodology

The premise for this study was based on the report by Douzgou et al^4^ and their survey formed the initial starting point for development. We then created an accessible questionnaire, designed for individuals with KBGS and their caregivers, to provide insight into how this condition affects a person’s daily life. The questionnaire was designed to capture information that may have been missed by healthcare professionals. It was co-produced by an international network of physicians, researchers and parents/carers of individuals with KBGS

Between December 2023 and June 2024, the study was predominantly advertised via the KBG Foundation, on their website and social media channels. Adults with KBGS were recruited to participate in the survey, either independently or with the support of family or carers. An adult was defined as a person over the age of 16 years due to this being a common delineator between paediatric and adult care. International collaborators from five other countries also shared the study with their patients, which was available translated into other languages. Participants were required to understand English, Spanish, French, Dutch, German or French to take part.

If an individual with KBGS was interested in participating, they were instructed to follow a URL link/QR code which directed them to an online participant information sheet. If participants wished to continue, they were asked to complete an online consent form. All consent data was collected via a web-based data capture system for research called Research Electronic Data Capture (REDCap). Participants were then asked to answer a series of questions relating to their KBGS (see supplementary information). Participants were able to save and return to their survey at any time to allow for breaks, if needed. Upon finalisation of data collection, outcome measurements were analysed using a combination of descriptive and simple frequency or percentage analysis.

## 3. Results

There were 104 responses to the survey. Of these, 10 were excluded from analysis because they did not complete the survey to the end, and three were excluded due to the participant being aged below 16years. This resulted in 91 responses for analysis. Some of these respondents did not answer every question , and the low response rate to individual questions was reflected upon during analysis.

Of the 91 responses, 66 were completed by caregivers and 25 were completed by the person with KBGS themselves. For the purpose of this paper the person with KBGS is referred to as the ‘participant’.

The ages of the participants in this survey ranged from 16-86years (median 22years, mean 24.8years). The ages were broken down into categories to enable further analysis of the data collected. There was a higher number of participants in the younger adult age range (see T able 1)

Forty-one participants were female, 44 male, and six did not respond to the question asking the gender of the participant.

Participants lived in the following countries: USA (n=25), United Kingdom (n=14), Spain (n=11), Netherlands (n=9), Denmark (n=7), Australia (n=3), France (n=3), Georgia (n=2), Germany (n=2), Italy (n=2), Norway (n=2), Belgium (n=1), Canada (n=1), Estonia (n=1), Portugal (n=1), South Africa (n=1) and this data was missing for six.

KBGS was diagnosed aged 6-9years for six participants, 10-17years old for 37, 18years or older for 41 while the age at diagnosis was missing for seven participants.

### 3.1 Everyday life

#### 3.1.1 Living arrangements

In total, 60 (66%) participants lived with their main caregiver, while nine (10%) were in supported living. Eleven (12%) lived independently, six (7%) reported there was ‘other’ arrangements and the data were missing for five (5%).

The data were analysed to look at the frequency of these living arrangements within each age category (see Figure 1). For 16-24year olds; 90% (n=47) lived with their main caregiver, 8% (n=4) in supported living and 2% (n=1) independently. For 25-34year olds the proportion of individuals living with their main caregiver decreased to 40% (n=8), with 15% (n=3) in supported living, 35% (n=7) independently, and 10% (n=2) ‘other’. The proportion of participants living at home with their main caregiver decreased further for 35-44year olds to 27% (n=3). This trend did not continue into the older age categories, however, there were less participants in these groups.

**Figure 1:**
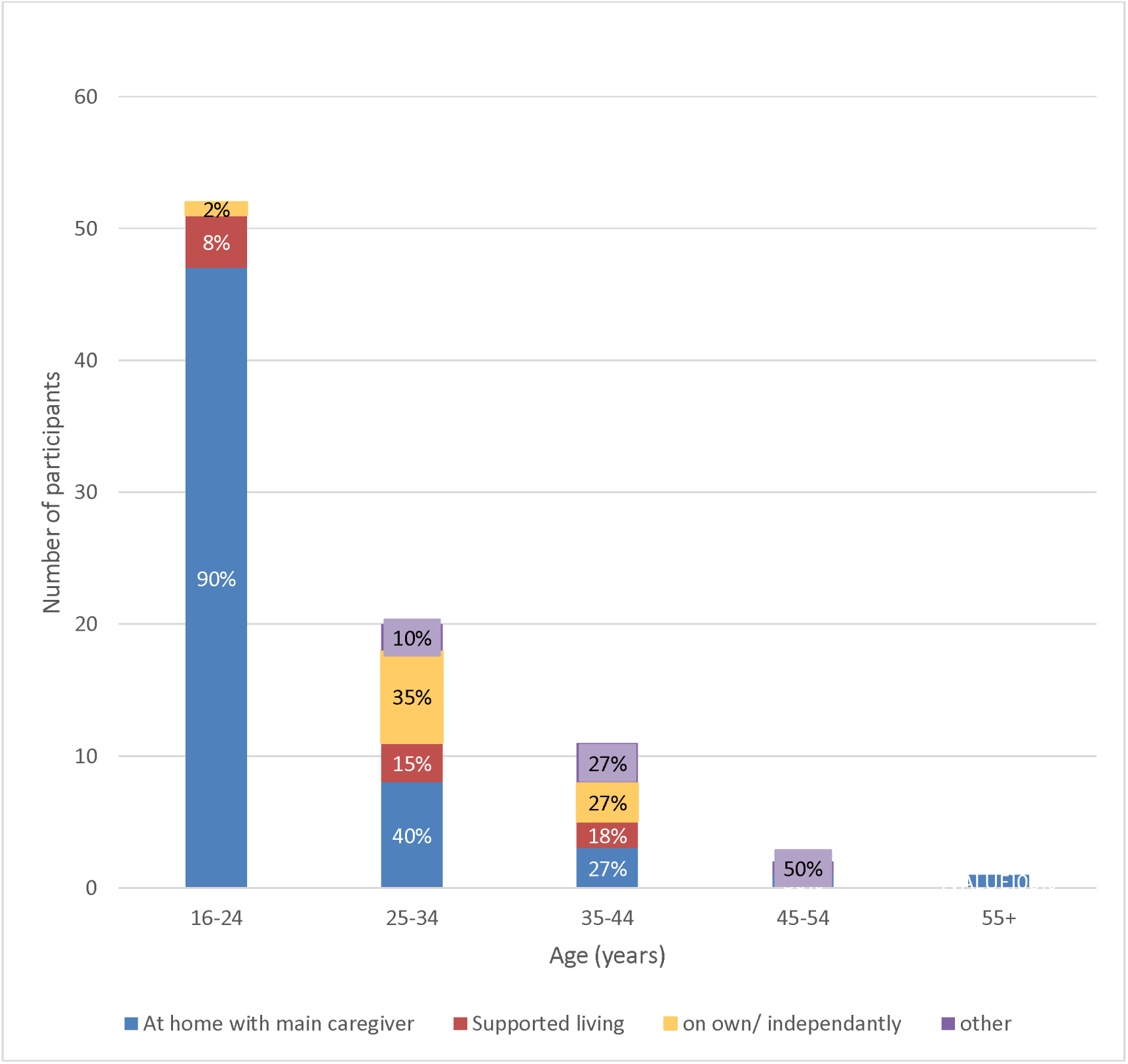
Chart comparing living arrangements across age categories of participants with KBGS

#### 3.1.2 Grocery shopping

In total, 47 (52%) participants were able to do their grocery shopping independently, and 34 (37%) reported they were not. The data were missing for 10 (11%) participants. The responses were analysed according to the age category of the participants (see figure 2). The proportion of individuals able to do their grocery shopping independently increased from 44% (n=23) in the 16-24 year olds, to 70% (n=14) in the 25-34year olds, to 73% (n=8) in the 35-44year olds, and 100% (n=2) in the 45-54year olds. The individual in the 55+ age group was unable to do their grocery shopping independently.

**Figure 2:**
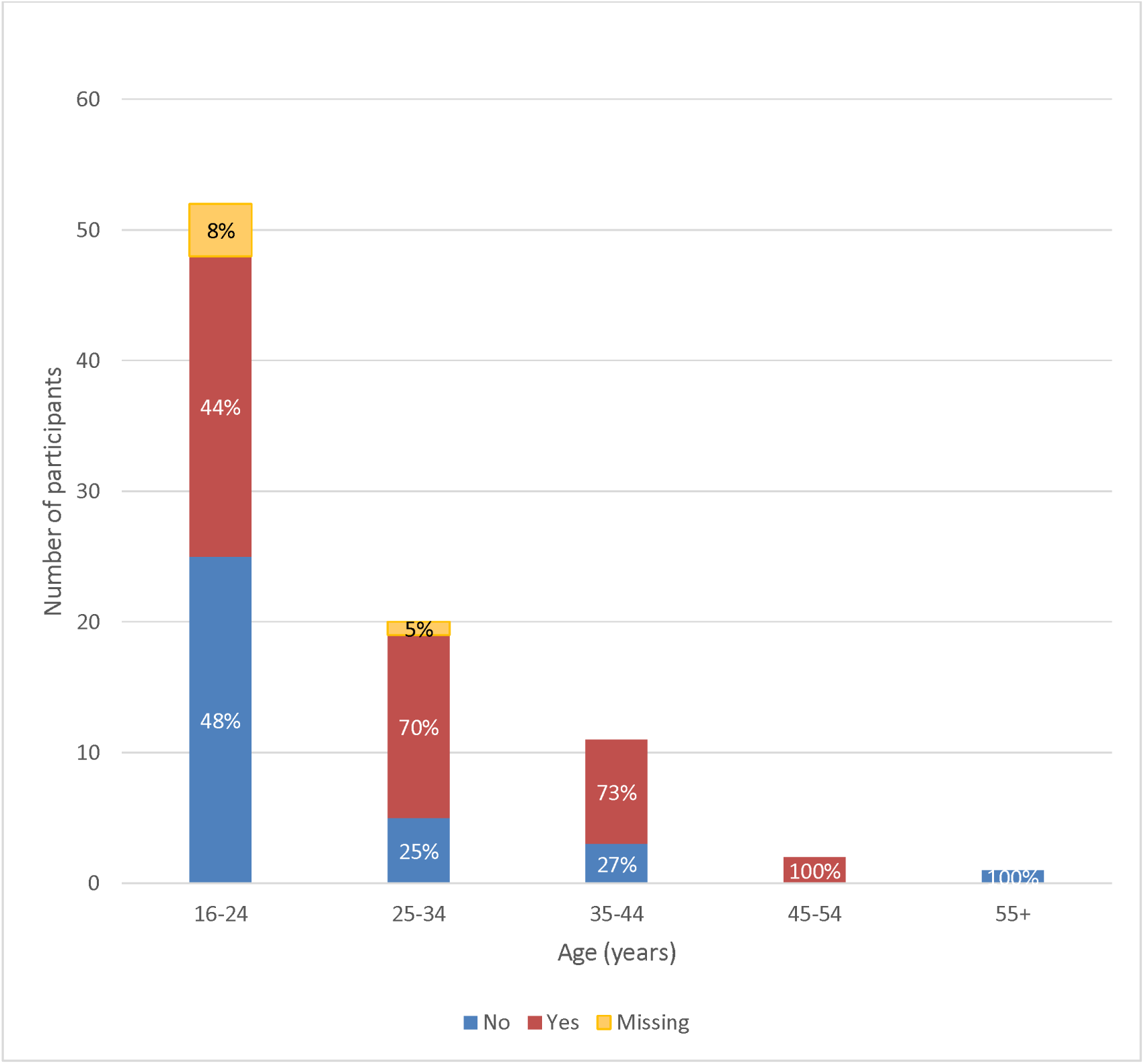
Chart comparing ability to do grocery shopping independently across age categories of participants with KBGS

#### 3.1.3 Driving

Seventy-one participants (78%) could not drive a car, 10 (11%) could and data for 10 (11%) was missing. Responses were analysed according to the age categories (see figure 3). The proportion able to drive a car increased from 2% (n=1) in the 16-24 year olds, to 20% (n=4) in the 25-35year olds, 36% (n=4) in the 35-44 year olds and 50% (n=1) in the 45-54year olds.

**Figure 3:**
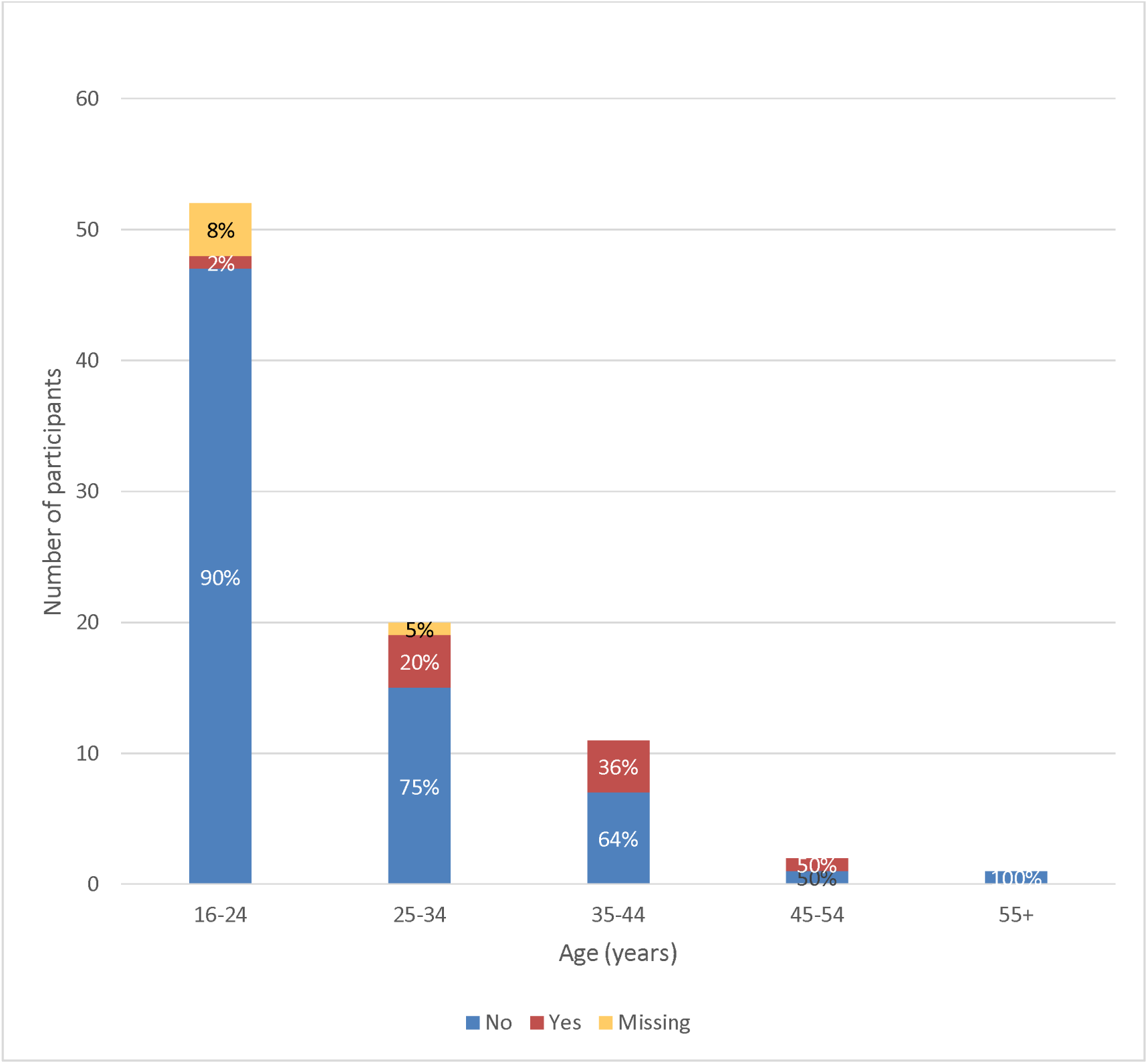
Chart comparing ability to drive across age categories of participants with KBGS

#### 3.1.4 Education

Eleven participants (12%) were currently enrolled in school, while 18 (20%) had completed high school or college. Twenty four (26%) described requiring a special educational setting or had some modifications to the curriculum. Sixteen (18%) had undertaken vocational training. One (1%) was currently attending university, one (1%) had obtained an undergraduate degree, and one (1%) had earned a master’s degree. Furthermore, two participants (2%) did not attend any educational institution. Data regarding educational attainment was missing for 50 participants (55%).

#### 3.1.5 Employment

Thirty participants (33%) were employed. There were a range of careers in the following job categories: Hospitality and food (n=6), Environment and Land (n=4), Retail and Sales (n=4), Home services (n=3), Social Care (n=3), Teaching and education (n=2), Administration (n=2), Animal care (n=1), Creative and media (n=1), Delivery and storage (n=1), Healthcare (n=1) and Sports and Leisure (n=1). One participant was employed at a sheltered workshop but did not specify the type of activities performed.

Twenty-nine reported whether the person with KBGS had any kind of support at work. Of these, 16 reported no specific support while three responded “yes” without further detail. Two mentioned having support persons in their team, one reported receiving support from teachers, three noted having one to one support and four described working in sheltered workshops for people with disabilities under adapted or protected conditions.

#### 3.1.6 Leisure

Fifty four respondents (60%) reported the person with KBGS maintained social activities. Of these, nine reported attending organised social activities/outings and nine participated in sports with one competing to an elite level. Six described involvement in creative arts; four music, one drama and one dance. Nine respondents went out with friends, going to the cinema (n=2), swimming (n=1) and sleepovers (n=1). Two respondents played computer games, three maintaining social connections through living and/or working with others and two through religious groups.

Two individuals reported their social activities were limited, and two enjoyed going out with family but do not socialise with friends. Two respondents described their challenges to accessing leisure activities; one did not like being in large groups of people alone and the other expressed they did not like change in routine or unpredictable events.

#### 3.1.7 Relationships

Seventy-four participants (81%) had never been in a romantic relationship. Of these, 25 expressed a desire for a relationship while another 25 stated they were not interested in romantic relationships.

Seventeen (19%) had been in a romantic relationship in the past, with nine of these currently in a relationship. Eleven (12%) reported currently had a long-term partner or spouse, while six (7%) had previously been in a long-term relationship.

When asked about sexual activity, 12 (13%) respondents confirmed they were sexually active, and 10 (11%) reported having children. Fifty-one indicated they were not sexually active, while three were unsure, three preferred not to answer, and 12 did not respond. Regarding self-stimulation, six respondents confirmed engaging in this behaviour, and nine indicated doing so with a partner.

### 3.2 Health issues

#### 3.2.1 Psychiatric and behavioural difficulties

Fifty-one (56%) reported psychiatric or behavioural difficulties affected their quality of life in childhood and 54 (59%) reported that such difficulties affected their quality of life in adulthood.

Fourty-four (48%) experienced anxiety while 40 (44%) and 37 (41%) had autism spectrum disorder and attention-deficit (hyperactive) disorder, respectively. Seventeen (19%) had experienced depression, and eleven (12%) reported recurring unwanted thoughts, ideas, or sensations (obsessions) that compelled them to act in certain ways. Seven (8%) experienced regression, marked by a loss of previously acquired skills, and four (4%) reported a sudden onset of reduced activity.

Additionally, one participant (1%) had schizophrenia, and one (1%) was diagnosed with borderline personality disorder.

High levels of frustration were reported in 34 (37%); 24 (26%) experienced episodes of rage, 23 (25%) displayed aggression towards themselves or others and 16 (18%) reported unspecified mood abnormalities. Sixteen (18%) reported sensory seeking behaviour and 10 (11%) sensory avoiding behaviour. It was an oversight that a question on intellectual disability was not specifically asked. However in the free text a range of presentations of intellectual disability were described, from mild to severe impairment and requiring support for all daily needs. Less common features included dyslexia, reported by two individuals.

Binge eating was reported in 14 (15%) and reduced eating or anorexia reported in 10 (11%). One respondent described experiencing hyperphagia leading to vomiting, along with challenges such as porn addiction and skin picking, which resulted in infections.

#### 3.2.2 Neurological

Twenty-eight (31%) reported neurological problems affecting their quality of life in childhood and 23 (25%) reported they were having an impact in adulthood.

A total of 27 (31%) participants reported experiencing seizures at some point. Of these, 14 (15%) reported that seizures were currently controlled by medication or ketogenic diet, while eight (9%) currently experienced treatment-resistant seizures. Six out of 27 had previously experienced seizures but no longer required medication. It was noted that of the 21 participants currently experiencing seizures, none of these individuals were living independently.

Although tics were reported in eleven participants (12%), none were diagnosed with Tourette’s syndrome. Three participants had tethered chord, two had anosmia, two experienced migraines, one had a ventriculo-peritoneal shunt due to hydrocephalus, one participant reported brain atrophy and one reported a stroke.

#### 3.2.3 Sleep

Nearly a quarter of participants (26%) reported that sleep problems negatively impacted their quality of life in childhood, with the same number stating these issues continued into adulthood. On the other hand, 22 (24%) did not experience any sleep-related issues. Difficulty falling asleep affected 25 (28%), with 11/25 requiring medication like melatonin to help initiate sleep. Similarly, 24 (26%) had trouble staying asleep, while 19 (21%) experienced an increased need for sleep. Suddenly falling asleep during the day was reported in eight participants. Additionally, five (5%) experienced sleep apnea, and four (4%) had restless leg syndrome.

#### 3.2.4 Gastrointestinal

A total of 21 participants (23%) reported that gastrointestinal problems negatively impacted their quality of life in childhood, while 26 (29%) indicated these issues persisted into adulthood.

Constipation was the most reported issue, affecting 26 participants (29%), followed by reflux, which affected 19 participants (21%). Other reported conditions included abdominal migraines (n=7), recurrent or cyclical vomiting (n=6), irritable bowel syndrome (n=5), chronic diarrhea (n=5), and inflammatory bowel disease (n=4). Less frequent conditions included diverticulitis/diverticulosis (n=2), eosinophilic esophagitis (n=2), gallstones (n=2), pancreatitis (n=1), and bowel obstruction (n=1).

#### 3.2.5 Cardiovascular

Cardiac problems affecting quality of life in childhood were reported in eight participants, with the same number indicating these issues persisted into adulthood. Heart valve problems were reported by 11 participants (12%), while hypertension was reported in eight participants (9%). Additional conditions included blood vessel dilatation (n=4), the need for heart medications (n=4), and cardiomyopathy (n=2). Additional cardiac features included sinus of Valsalva aneurysm (n=1), mild aortic dilatation (n=2) and moderate aortic insufficiency with valve sclerosis (n=1), dextrocardia (n=1), mitral valve displacement (n=1), dilated coronary sinus (n=1), and a miniscule heart hole (n=1). Prolonged QT syndrome, ectopic atrial rhythm, and pericarditis were each reported by one participant. Four participants reported heart surgeries: one for a bicuspid aortic valve with dilatation of the aortic root, one for atrioventricular septal defect, one for ventricular septal defect, and one for primum atrial septal defect with mitral valve cleft repair.

#### 3.2.6 Musculoskeletal

Twenty-six (29%) had scoliosis while 25 (27%) had lordosis or kyphosis. Hip problems were noted by 14 (15%), arthritis by seven (8%), and osteoporosis by five (5%). Additional musculoskeletal issues included foot deformities described by seven participants. One participant underwent surgery for Grisel’s syndrome, and another for the removal of a FGF23 -producing tumour of the scapula.

#### 3.2.7 Hearing and vision

Thirty-four (37%) of respondents reported hearing problems affected their quality of life in childhood and 32 (35%) reported they are still affecting their quality of life in adulthood. Two participants had cochlear implants. One participant had enlarged vestibular aqueducts requiring hearing aids. One participant reported a tympanic homograft and tympano ossicular graft and another a tympanoplasty. Two individuals reported cholesteatoma, with another reporting chronic ear inflammation. Three respondents had operations on the adenoids, two of whom had insertion or widening of ear ventilation tubes.

Thirty-three (36%) participants reported visual difficulties. Two respondents reported Salzmann’s nodules on the cornea. One participant had keratoconus corneal dystrophy, and another had microcornea . One respondent described a hole in the eye requiring an emergency corneal transplant. Two respondents had strabismus, one of whom had lens implants.

#### 3.2.8 Dental

A total of 28 (31%) exhibited misshapen teeth, while 20 (22%) encountered issues with dental enamel. Additionally, 17 (19%) reported problems with the palate, 17 (19%) experienced dental decay, and another 17 (19%) had an excess of teeth. Furthermore, five (5%) reported experiencing crumbling teeth. Two participants indicated that their deciduous teeth did not fall out, and five reported the absence of certain teeth.

#### 3.2.9 Other

Additional features included keloid scars (n=7) and abnormal wound healing (n=7). Two participants had hypothyroidism, while one reported further unspecified thyroid problems. Kidney problems were reported in two and included duplex kidneys or one small kidney. One was reported to have glucose-6-phosphate dehydrogenase deficiency, Gilbert’s disease, and lymphatic malformations.

One participant had a cleft palate. One participant had surgery for choanal atresia. One participant experienced velopharyngeal insufficiency, with another describing a nasal voice. Lastly, one participant reported undergoing surgery for a small cell tumor of the ovary, and one respondent had a pituitary cyst along with polycystic ovary syndrome.

## 4. Discussion

This study found independent living, grocery shopping and driving skills, were acquired by some adults with KBGS and that the prevalence of skill attainment was mostly highest in the 35-44 age group. This is important information for individuals with KBGS, their families and genetic counsellors when planning for the future. However considering the cross sectional study design and difference in absolute numbers of each age category, it is difficult to conclude the timescale of skill acquisition.

In this cohort 33% reported employment, comparable to the 28% in our previous physician reported data, and in contrast to only 8% in the literature group from the same study^5^. We recognise that socioeconomic circumstances are likely to be a significant factor in an individual’s ability to live and work, and that furthermore there is potential bias, whereby those with more support will have been more able to respond to this survey.

Some adults with KBGS will have only received genetic testing due to having children with more complex problems, and given some of the difficulties experienced with this condition can be mild and non-specific there can be delays in diagnosis^6^. Most individuals with KBGS have a mild intellectual disability but in a small proportion this can vary from no intellectual disabilities to severe^3^. The responses demonstrate that high educational attainment is possible in KBGS and it is plausible that there are further very mildly affected undiagnosed individuals in this category.

None of the participants aged 45+ in this data set lived independently, which could correlate with a testing bias. Given the changes in availability of genetic testing, it is likely that there are unidentified older adults with KBGS who will never undergo genetic testing, and that the older adults in this group have been diagnosed due to having a more significant set of problems. These figures may change in the future as the diagnosed children grow up. We hope the impact of an early diagnosis with appropriate support and interventions may also result in some further positive shift of these figures. Using the information from our results we have considered the types of support that could be needed, along with possible early interventions to maintain long-term health and wellbeing outcomes.

Given the high burden of psychiatric and behavioural difficulties we recommend that adults with KBGS are provided psychosocial support to optimise their mental health. This may include onward referral to psychiatry for assessment and treatment. We recognise best practice is a holistic approach, where support can be provided by wider members of multidisciplinary teams, such as the positive behavioural approach, in line with NICE guidelines for behaviours that challenge^7^.

Reasonable adjustments to work and education should be made to ensure individuals have the best chance of achieving their potential. Sensory needs assessments could inform these when indicated. Annual hearing and vision checks should be aimed for annually, due to the high rates of hearing and visual problems, which can also be contributing factors to psychiatric and behavioural difficulties, and sensory seeking/avoiding behaviours.

This study found 21 (23%) participants were currently experiencing seizures, compared to our previous study where 28% of the adults with KBGS had epilepsy^5^. For the participants in this study, none of those who currently experienced seizures were found to be living independently. Although, other factors, such as cognitive function and social support could also influence independence, this observation suggests that seizures significantly impact daily living capabilities, limiting independence and necessitating additional support and supervision.

Unfortunately, a question around the impact of musculoskeletal problems on quality of life in adulthood was not asked in this survey, but we have inferred the high proportion of skeletal issues to indicate these are having an impact and would advise optimising bone health, maintaining healthy physical activity, and appropriate investigation and treatment of painful joints.

Congenital heart disease and dilatation of the aorta were reported at higher rates than expected from the general population. In our previous study, we also identified cardiovascular abnormalities relating to the ascending aorta (n=2) and/or aortic valve (n=3)^5^. We acknowledge some individuals are likely to be reported in both our papers. However, these associations have also been reported in the wider literature and warrant further investigation^8,9^. Based on this, we recommend consideration of cardiac ECHO including a measurement of the aortic diameter, at least once in adulthood (or more regularly if abnormalities are identified requiring monitoring or if symptomatic).

We found a high burden of gastrointestinal issues into adulthood, including cyclical vomiting and abdominal migraine which has also been reported in the literature^10^. Constipation and reflux were identified in our previous study and again were common complaints in this cohort. This information could be used to raise awareness for practitioners who should treat as per condition on a case by case basis. See figure 4 for our recommendations for healthcare interventions for adults with KBGS.

**Figure 4:**
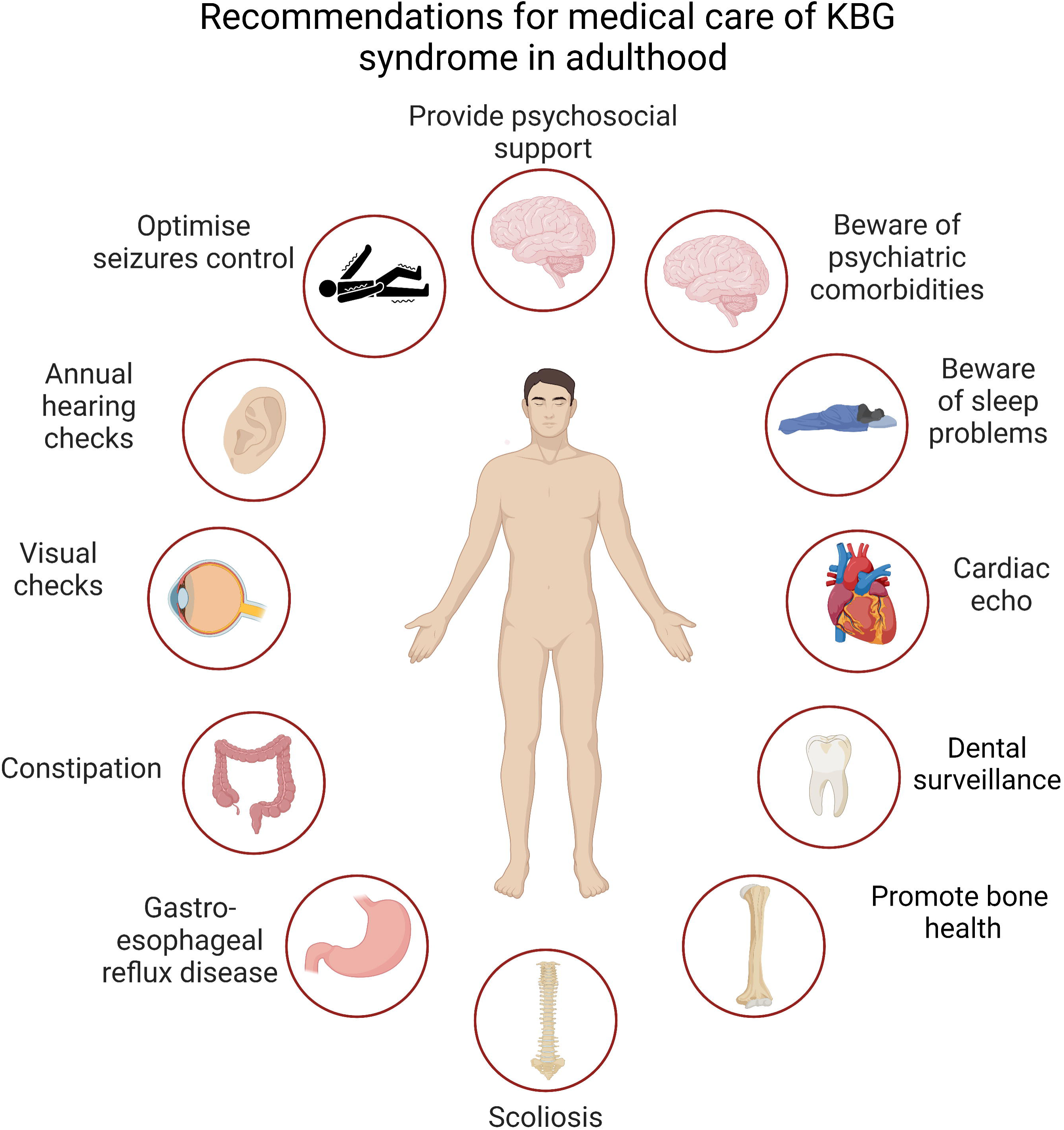
Image summarising the recommendations for medical care of KBGS in adulthood

There are limitations to this study that require consideration when making wider conclusions about the outcomes of KBGS. Sixty-six (73%) of surveys were completed by the caregiver rather than the individual with KBGS which could infer a level of intellectual disability which would make completing such a survey difficult or unachievable. We note that educational attainment responses were missing in 50 cases – this could be because of limited educational attainment or because of problems understanding the questions. If this sort of survey is used in the future the specific questions regarding intellectual disability and educational attainment should be revised, and ideally tested first with a small group, to ensure it provides a better response rate and a better reflection of intellectual disability. It is also possible there were questions that respondents felt embarrassed or uncomfortable answering, for example around sexual relationships, and further refining these questions through consultation with individuals with KBGS would be beneficial.

This method of using adult/caregiver responses is vulnerable to recall bias and subjectivity, along with potential challenges arising from limited medical knowledge of the respondents. We found during analysis we were not able to ascertain in all responses, whether an individual’s medical problem or surgery occurred in childhood or adulthood, and that our questions could have been misunderstood. There were also inconsistencies in respondents reporting whether conditions were autoimmune disorders, which has limited comparisons we have been able to make with our previous paper in this area. Due to the paucity of data we have reported the majority of our findings, and acknowledge that some of the rates of these will not be out of keeping with the general population. However, the strengths of this method of online questionnaire participation have included our success in recruitment of a large cohort across a number of countries, and identification of implications for clinical management along with areas for further exploration. The method of our patient/carer questionnaire is likely to be of interest to researchers in a variety of other settings in the field of rare genetic syndromes.

## 5. Conclusion

Our study has clearly shown the wide range of education and employment that can be achieved by individuals with KBGS, along with social activities and relationships that they can have. It can therefore be concluded that since KBGS can affect people’s lives in such varied ways, the diagnosis alone should not influence an individual’s future aspirations. There are, however, high rates of comorbidities, which continue to affect people’s quality of life into adulthood. Support will need to be tailored to the individual’s needs, and most likely will require the input of professionals from multiple specialities in order to optimise their health and wellbeing into adulthood. This study, through utilising the adult’s and caregiver’s perspective, provides valuable insights into the real world needs of adults with this condition. This knowledge can be used to enhance counselling, clinical decision making, care coordination and inform the development of holistic evidence-based guidelines.

**Table 1.** Age of participants with KBGS.

| Age | Number of participants | Percent |
| --- | --- | --- |
| 16-24 | 52 | 57.1 |
| 25-34 | 20 | 22 |
| 35-44 | 11 | 12.1 |
| 45-54 | 2 | 2.2 |
| 55+ | 1 | 1.1 |
| Missing | 5 | 5.5 |
| Total | 91 | 100 |

## Acknowledgements

We thank the KBG Foundation for their support and we would like to thank the individuals with KBG Syndrome and their families and carers for taking part in this study. Extended thanks and gratitude goes to Paige Fumo Fox and Annette Maughan, for their time and participation in the development of this research and their valuable insights as Experts by Experience. We thank Victor Mulerno for his assistance in translating the study documents into Spanish and for his useful comments.

## Funding Statement

Allan Bayat is funded by a BRIDGE -Translational Excellence Programme grant funded by the Novo Nordisk Foundation, grant agreement number: NNF20SA0064340.

Karen Low is funded by an NIHR Doctoral research fellowship award: NIHR302303. The views expressed in this publication are those of the author(s) and not necessarily those of the NIHR, (Partner Name), NHS or the UK Department of Health and Social Care.

Nuria C Bramswig is a member of the European Reference Network on Rare Congenital Malformations and Rare Intellectual Disability ERN-ITHACA. ERN ITHACA is funded by the European Union, under the Grant Agreement nr. 101156387.

## Ethics Declaration

We confirm that we have read the Journal’s position on issues involved in ethical publication and affirm that this report is consistent with those guidelines. The study complies with the principles laid out in the Declaration of Helsinki. The study was approved by the faculty of health research ethics committee at the University of Bristol/UK, in November 2023 (ref 16025). Written informed consent was collected from all respondents prior to completing the questionnaire, using research protocols approved by the ethics committee at the University of Bristol. The data gathered via the questionnaire was designed to allow anonymity and confidentiality.

## Author contribution

TBC

## Declaration of interests

The authors declare no conflict of interests.

## Data availability

De-identified data will be made available to those eligible per request to the corresponding author.

## Legends

**Supplementary Figure 1:** Specialities following up KBGS in adulthood

**Supplementary File 1:** Copy of questionnaire ‘Life Beyond Childhood: Understanding Adult Aspects of KBG Syndrome’

## Notes

### Competing Interest Statement

The authors have declared no competing interest.

### Author Declarations

The study was approved by the faculty of health research ethics committee at the University of Bristol/UK, in November 2023 (ref 16025). Written informed consent was collected from all respondents prior to completing the questionnaire, using research protocols approved by the ethics committee at the University of Bristol. The data gathered via the questionnaire was designed to allow anonymity and confidentiality.

