## Supplementary file 1 for "Life beyond childhood: insight into the lived experience of 91 adults with KBG syndrome through an online patient/caregiver reported co-produced questionnaire"

### Life Beyond Childhood: Understand Adult Aspects of KBG Syndrome

Consent ID

__________________________________

We are collecting information about individuals (>16-years-old) with KBG syndrome. We aim to develop guidelines for the management of their specific needs. We would be grateful if you would fill in the following questionnaire. The questionnaire should take approximately 10 minutes to complete. You can also save and return later if you want to take a break. None of these questions are required but your complete responses will be of great help. Questions are worded so that either the person with KBG syndrome or a caregiver may complete it. There should be only one respondent from each family. If there is more than one person in the family with KBG syndrome, a new questionnaire should be completed for each person. Your submission is anonymous, but if you want to include your email in the "Final Comments" Question, at the end of this questionnaire, we will be able to contact you in case we have any questions. Please review your answers before submitting them to ensure they are as complete and correct as possible. Feel free to email the study team on

#### GENERAL QUESTIONS

| | |
|---|---|
| Are you a caregiver or a patient (individual with KBG syndrome)? | ◯ Patient<br>◯ Caregiver |
| Please explain your answer:<br>For example, are you a parent? | __________________________________ |
| How old is the person with KBG syndrome? | __________________________________ |
| In which country does the person with KBG syndrome live? | __________________________________ |
| Where does the person with KBG syndrome live? | ◯ At home with their main caregiver full-time (e.g., with family)<br>◯ In supported living services (a living arrangement with support from carers for everyday tasks)<br>◯ On their own / Independently<br>◯ Other |
| Please explain your answer: | __________________________________ |
| At what age was the person with KBG syndrome diagnosed? | ◯ Prenatally (before birth)<br>◯ Newborn/Infancy (less than 1-year of age)<br>◯ 1-5 years old<br>◯ 6-9 years old<br>◯ 10-17 years old<br>◯ 18-years or older<br>◯ I do not know |
| Has the KBG diagnosis been confirmed with genetic testing? | ◯ Yes<br>◯ No<br>◯ I do not know<br>◯ Test in progress / awaiting results<br>◯ Other |
| Would you like to give us more information?<br><br>For example, if known to you, you can write here the specific gene change. If possible, please add the genetic change (mutation) as the following example: NM_013275.5, c.5841del; p.Val1948Leufs*15. | __________________________________ |
| What is the gender of the person with KBG syndrome? | ◯ Female<br>◯ Male<br>◯ Other<br>◯ Prefer not to answer |
| Please explain your answer: | __________________________________ |
| What is the person with KBG syndrome's current height in centimeters (cm)? | __________________________________ |

| | |
|---|---|
| Did the person with KBG syndrome have any of the following medical problems during childhood? | ☐ Behavioral/psychiatric problems<br>☐ Gastrointestinal problems<br>☐ Neurological problems (e.g., seizures)<br>☐ Sleep problems<br>☐ Heart problems<br>☐ Hearing problems<br>☐ Other<br>☐ None of the above |
| Would you like to explain your answer(s)? | __________________________________ |
| Did any of the medical problems impact their quality of life in childhood?<br>Which problems had an impact? | ☐ Behavioral/psychiatric problems<br>☐ Gastrointestinal problems<br>☐ Neurological problems<br>☐ Sleep problems<br>☐ Heart problems<br>☐ Hearing problems<br>☐ Other<br>☐ None of the above |
| Are any medical problems still impacting their quality of life in adulthood?<br>Which problems have an impact? | ☐ Behavioral/psychiatric problems<br>☐ Gastrointestinal problems<br>☐ Neurological problems<br>☐ Sleep problems<br>☐ Heart problems<br>☐ Hearing problems<br>☐ Other<br>☐ None of the above |

#### GASTROINTESTINAL PROBLEMS

If the person with KBG syndrome currently has or has had gastrointestinal problems in adulthood, can you find them in the list below?

- ☐ Reflux
- ☐ Constipation
- ☐ Abdominal migraine
- ☐ Blood in stool
- ☐ Chronic diarrhea
- ☐ Diverticulitis / Diverticulosis
- ☐ Eosinophilic esophagitis
- ☐ Esophageal varices
- ☐ Gallstones
- ☐ Hirschsprung disease
- ☐ Inflammatory Bowel Disease (i.e., Crohn's, Ulcerative Colitis)
- ☐ Irritable Bowel Disease/Syndrome
- ☐ Liver fibrosis
- ☐ Liver problems
- ☐ Malrotation (abnormality of the bowel)
- ☐ Pancreatitis
- ☐ Recurrent/cyclic vomiting
- ☐ Small Bowel Obstruction
- ☐ Unsure
- ☐ Other
- ☐ None

Would you like to explain your answer(s)? ______________________________

Please provide further detail about the liver problems: ______________________________

How frequently are stools coming typically in a week? ______________________________

What is the person with KBG syndrome using to manage their constipation?

- ☐ A specific diet
- ☐ Medications
- ☐ Other

Please explain your answer:
For example, what kind of diet? ______________________________

Is a specific management / treatment more helpful than others? If so, which one? ______________________________

#### SKIN & PHYSICAL HEALTH PROBLEMS

If the person with KBG syndrome currently has or has had skin problems in adulthood, can you find them in the list below?

- ☐ Heavy or hypertrophic (enlargement of an organ/tissue) scars that can occur after an injury to the skin, also known as keloids
- ☐ Abnormal scarring of other type
- ☐ Poor wound healing
- ☐ Increased skin infections
- ☐ In growing or infections of their fingernails or toenails that require treatment with medication
- ☐ Very dry skin
- ☐ Eczema
- ☐ A yellowish or greenish pigmentation of the skin and, sometimes, of the white of the eyes, also known as jaundice
- ☐ Swelling of hands and feet
- ☐ Vitiligo
- ☐ Acne
- ☐ Other
- ☐ Unsure
- ☐ No problems

Would you like to explain your answer(s)?

__________________________________

Did the person with KBG syndrome require treatment with growth hormones in childhood?

- ◯ Yes
- ◯ No
- ◯ I don't know
- ◯ Prefer not to answer

Does the person with KBG syndrome have any of the following?

- ☐ Scoliosis
- ☐ Curve of the spine (lordosis/kyphosis)
- ☐ Hip problems
- ☐ Arthritis
- ☐ Osteoporosis
- ☐ Other

Please explain your answer:

__________________________________

Does the person with KBG syndrome have any of the following?

- ☐ Heart valve problems
- ☐ Heart muscle weakness (cardiomyopathy)
- ☐ High blood pressure
- ☐ Widening of the blood vessels (dilatation)
- ☐ Requires heart medications
- ☐ Other

Please explain your answer:

__________________________________

Does the person with KBG syndrome have any of the following?

- ☐ Seizures currently - medication controlled, including ketogenic diet
- ☐ Seizures currently - difficult to treat, not currently under control
- ☐ Seizures previously - no longer on medication
- ☐ Other

What medications?

__________________________________

| | |
|---|---|
| Please explain your answer: | __________________________________ |
| Does the person with KBG syndrome have any of the following? | ☐ Problems with dental enamel<br>☐ Dental decay<br>☐ Too many teeth<br>☐ Crumbling teeth<br>☐ Misshapen teeth<br>☐ Problems with the palate<br>☐ Other |
| Please explain your answer: | __________________________________ |

#### SLEEP PROBLEMS

If the person with KBG syndrome currently has or has had sleep problems in adulthood, can you find them in the list below?

- ☐ No sleep problems
- ☐ Difficulty falling asleep
- ☐ Difficulty staying asleep/awaking frequently during the night
- ☐ Increased need for sleep
- ☐ Falling asleep suddenly during the day / Narcolepsy
- ☐ Breathing repeatedly stops and starts during sleep, or the person snores loudly and feels tired even after a full night's sleep. May also be known as Sleep Apnea
- ☐ Unpleasant or uncomfortable sensations in the legs and an irresistible urge to move them. Symptoms commonly occur in the late afternoon or evening hours and are often most severe at night when a person is resting, such as sitting or lying in bed. Can also be known as Restless Legs Syndrome or Periodic Limb Movement Disorder
- ☐ Requires medication such as melatonin to initiate sleep
- ☐ Other
- ☐ Unsure

Please explain your answer: ______________________________

Which one of these options best describes the problem 'increased need for sleeping'?

- ☐ Increased night time sleeping
- ☐ Increased day time sleeping
- ☐ Increased need to sleep immediately after lunch
- ☐ Mood change if not allowed to sleep
- ☐ Other

Please explain your answer: ______________________________

#### BEHAVIOURAL / PSYCHIATRIC PROBLEMS

If the person with KBG syndrome currently has or has had behavioral/psychiatric problems in adulthood, can you find them from the list below?

- ☐ Aggression toward others or self
- ☐ High levels of frustration
- ☐ Episodes of rage
- ☐ Anxiety
- ☐ Autism Spectrum Disorder
- ☐ Depression
- ☐ Schizophrenia
- ☐ Tics
- ☐ Tourette syndrome
- ☐ Difficulty maintaining attention, hyperactivity and impulsive behaviour. May also be known as attention deficit hyperactivity disorder (ADHD)
- ☐ Limited interests and repetition (routine/words/order of doing things)
- ☐ Loss of interest in things that used to interest them
- ☐ Loss of skills that they used to be able to perform (regression)
- ☐ Mood abnormalities or disorder/sudden mood changes
- ☐ Recurring, unwanted thoughts, ideas or sensations (obsessions) that make them feel driven to do something repetitively (compulsions), also known as obsessive compulsive disorder (OCD)
- ☐ Self-harm/Aggression toward themselves
- ☐ Sudden onset of reduced activity
- ☐ Sensory seeking behavior (including self-stimulatory behavior)
- ☐ Sensory avoiding behavior
- ☐ Other
- ☐ Unsure
- ☐ No problems

---

Please explain your answer:

__________________________________

---

When did the behavioural/psychiatric problems start?

- ☐ During childhood
- ☐ During puberty
- ☐ In adulthood

---

If you chose more than one option and the problems started at different times, please could you specify?

__________________________________

---

How were the behavioural/psychiatric problems managed?

- ☐ Change of environment
- ☐ Psychotherapy
- ☐ Medications

---

If you chose more than one option and each problem was managed differently, please could you specify?

__________________________________

---

Was a specific management more helpful than others and if so, which one and for which behavioral problem?

__________________________________

---

Which of the behavioural problems are influencing the life of the person with KBG syndrome the most?

__________________________________

#### OTHER PROBLEMS

Are there other problems that the adult person with KBG syndrome has?

- ☐ Binge eating
- ☐ Reduced eating or anorexia
- ☐ Cancer
- ☐ Decreased mobility/any difficulties with moving around
- ☐ Autoimmune conditions
- ☐ Diabetes
- ☐ Hearing difficulties or loss
- ☐ High pain threshold
- ☐ Hypersensitivity to noise and crowded places
- ☐ Hypertension or other problems with the blood vessels and the heart
- ☐ Menopause
- ☐ Kidney illness
- ☐ Vision difficulties or loss
- ☐ Other

Please explain your answer: ______________________________

How old were you when the menopause started? ______________________________

#### SURGERY

| | |
|---|---|
| Has the person with KBG syndrome had any surgery during adulthood? | ◯ Yes<br>◯ No |
| What was the original problem? | ____________________ |
| How many times has the person with KBG syndrome had surgery? | ____________________ |
| What type of surgery did the person with KBG syndrome have? | ____________________ |
| Did the surgery resolve the issue? | ____________________ |
| Do you have any other comments about surgery in the person with KBG syndrome? | ____________________ |

#### EVERYDAY LIFE

What level of education did the person with KBG syndrome achieve? Please provide detail.

__________________________________________

Does the person with KBG syndrome work?

◯ Yes
◯ No

Please specify the type of work:

__________________________________

Does the person with KBG syndrome have any kind of support at work?

__________________________________

Does the person with KBG syndrome maintain social activities?

◯ Yes
◯ No

Please explain your answer:

__________________________________

Does the person with KBG syndrome drive a car?

◯ Yes
◯ No

Is the person with KBG syndrome able to take transportation independently?

◯ Yes
◯ No

Is the person with KBG syndrome able to do grocery shopping independently?

◯ Yes
◯ No

#### ROMANTIC RELATIONSHIPS

| | |
|---|---|
| Please select all that apply. The person with KBG syndrome has: | ☐ Had a romantic relationship in the past<br>☐ Is currently in a romantic relationship<br>☐ Has a long-term partner or spouse<br>☐ Previously had a long-term relationship<br>☐ Would like a relationship but has never had one<br>☐ Is not interested in a romantic relationship |
| A question caregivers and parents often ask is whether individuals with KBG syndrome have been sexually active. We would be grateful if you would consider answering whether the adult KBG individual you are/know has been sexually active? | ◯ Yes<br>◯ They have a child/children<br>◯ No<br>◯ I do not know<br>◯ Prefer not to answer |
| Please could you specify? | ☐ Stimulation of one's own genitalia (masturbation)<br>☐ With a partner<br>☐ Other |
| Please explain your answer: | __________________________________ |
| Are there other problems that the person with KBG syndrome has that you would like us to know about? | __________________________________ |

#### MEDICAL FOLLOW-UP

| | |
|---|---|
| Who is following up the medical issues of the person with KBG syndrome? | ☐ The general practitioner<br>☐ A specialist in the hospital<br>☐ A reference centre where I/he/she are seen by many specialists<br>☐ Other<br>☐ No one. I/he/she has not seen any doctor in adult life |
| Please could you specify which specialty? | __________________________________ |
| Please could you specify which centre? | __________________________________ |
| Please explain your answer: | __________________________________ |
| How often does the person with KBG syndrome have health checks? | ◯ Once a year<br>◯ Twice a year<br>◯ Once every 3 years<br>◯ Once every 5 years<br>◯ Only if there are specific concerns<br>◯ Other |
| Please explain your answer: | __________________________________ |
| In the past 2 years, has the person with KBG syndrome needed to attend hospital for an unplanned reason or emergency? | ◯ Yes<br>◯ No |
| Please give the number of occasions and reason(s) for attendance: | __________________________________ |

#### FINAL COMMENTS

You have now completed the survey!

Is there anything else you would like to share? Any final comments? __________________________________
