## supplementary figure 1 for "Life beyond childhood: insight into the lived experience of 91 adults with KBG syndrome through an online patient/caregiver reported co-produced questionnaire"

**Supplementary Figure 1: Follow up in adulthood**

Respondents were asked who follows up the medical issues of the person with KBGS. Forty six reported follow up from their GP, 39 from a specialist in hospital, 14 from a reference centre where they are seen by many specialists, 10 responses were ‘other’ and 8 reported no-one/not seen a doctor in adult life.

The respondents were asked to describe which specialities they were followed up by and there were a wide range reported (see supplementary figure 1)

Supplementary Figure 1: Specialities following up KBGS in adulthood

|  | Number of participants |
| --- | --- |
| Neurology | 13 |
| Cardiology | 10 |
| Opthalmology/Optometry | 8 |
| ENT | 7 |
| Psychiatry/psychology | 7 |
| Audiology | 5 |
| Gastroenterology | 5 |
| Gentics | 2 |
| Dental | 2 |
| Dermatology | 2 |
| Endocrinology | 2 |
| Orthopedics | 2 |
| Paediatrics | 2 |
| Respiratory | 2 |
| Rheumatology | 2 |
| Cardiac surgery | 1 |
| Neurosurgery | 1 |
| Gynaecology | 1 |
| Allergist | 1 |
| Rare diseases/metabolic | 1 |
| Haematology | 1 |
| Internal medicine | 1 |
| Rehabilitation | 1 |
| Complex care | 1 |
| Physiotherapy | 2 |
| Occupational therapy | 1 |
| Speech and Language Therapy | 1 |
| Art therapy | 1 |

There was a wide range of complexity in the care of individuals, with some seeing multiple specialities and others having no health care needs in adulthood.

The frequency of follow up ranged from twice a year (23), once a year (21), only if there are specific concerns (15), once every 3 years (4), once every 5 years (3) and ‘other’ (9). The majority of respondents reporting ‘other’ (8/9) went on to describe more frequent follow up than twice a year, with 1 giving no further details.
